# Assessing Contributing and Mediating Factors of Telemedicine on Burnout

**DOI:** 10.1101/2023.05.08.23289673

**Authors:** Valerie Boksa, Priyadarshini Pennathur

**Author notes:** Corresponding Author: Valerie Boksa. There are no competing interests in this submission. Ethics approval was granted by the University of Iowa Institutional Review Board on 8/15/2022.

## Abstract

**Objective:** The prevalence of burnout among healthcare providers has been steadily increasing, with a call to action issued in 2019. Immediately following this call to action, the COVID-19 pandemic drastically changed demand. Use of telemedicine expanded in response to COVID-19 and changed the experience of care delivery for healthcare providers. The impact of telemedicine use during COVID-19 on the provider well-being is less well known. This study aims to assess the prevalence of burnout in providers who used telemedicine and to better understand how specific factors of telemedicine can impact workplace stress.

**Methods:** Providers in urgent care clinics were invited to participate in a burnout assessment survey using the Maslach Burnout Inventory questionnaire. The prevalence of burnout, burnout profiles, and correlations were analyzed in the resulting data. Follow-up interviews provided further insight on contributing and mediating factors of telemedicine on provider burnout.

**Results:** The findings from this study provide technology- and organizational-level recommendations to prevent increased risk of burnout among telemedicine providers. The classification of contributing and mediating factors also provides a framework for understanding the risks that this technology can pose to workplace stress. Future research recommendations to better quantify the relationship between burnout and telemedicine use and to effectively design intervention and implementation strategies are discussed.

**Public Interest Summary:** Considering the high rates of burnout in the healthcare industry prior to the pandemic, the severe demands the COVID-19 pandemic had on healthcare workers, and the drastic changes in workflow due to the widespread adoption of telemedicine, it is important to assess current levels of provider burnout and to collect information from frontline clinicians on how telemedicine impacts workplace stress. A survey was administered to assess burnout in healthcare workers who provided care via telemedicine. The interviews provided additional insight on how telemedicine affected workplace stress. Survey results showed that 25% of the respondents reported one or more manifestations of burnout; and there was a correlation between personal accomplishment scores and reported months of telemedicine use. Findings from the interviews and review of literature identified what design and use characteristics of telemedicine contributed to and/or alleviated burnout. Results address how organizations can best support their employees who administer care via telemedicine and guide researchers with direction for future studies.

## Introduction

The COVID-19 pandemic significantly altered the demand for telemedicine. With a high risk of illness, numerous organizations used telemedicine to continue providing remote care for patients. (1–4). In the last week of March, 2020 alone, some healthcare centers experienced a 154% increase in telemedicine visits compared to the previous year (1). Additionally, within the first two weeks of stay-at-home orders, some US clinics converted up to 86% of appointments to virtual formats (5). Telemedicine has long been available in healthcare, but previously many regulations were in place (6). However, the pandemic compelled hospitals to conserve resources and protect staff, resulting in a relaxation of regulations on location, platforms, and reimbursement for telemedicine use (5,7).

COVID-19 also substantially altered the workflow of healthcare providers and introduced significant stress and workload. Staff ratios and workload were impacted by a rise in the number of patients requiring care and a decline in the number of available clinicians. (8,9). Restrictions in organizational resources affected worker pay and access to personal protective equipment (8,10,11). Some employees also reported emotional distress about the risk of spreading infection to loved ones from occupational exposures (10). Burnout of healthcare workers, a measure of emotional exhaustion, depersonalization, and personal accomplishment (12), which was on the rise **<u>prior</u>** to the COVID-19 pandemic only worsened. Nationally, the prevalence of burnout in physicians is documented to range from 40% to 54% and nursing burnout rates vary between 35-45% (13). U.S. physician burnout prevalence has been increasing with 45.5%, 54.4%, and 43.9% for the years 2011, 2014, and 2017 respectively (2). A more recent nationwide assessment of burnout during the pandemic, by Shanafelt et al., focusing on physicians in all specialties suggest that manifestations of burnout symptoms in physicians are as high as 62% (2). But it is unclear whether a subset of these physicians had telemedicine experience given that the focus of their study was on all specialties. Shanafelt et al.’s evidence on burnout provides valuable information on the alarming rise of burnout, but what is unclear is how telemedicine contributes to or mitigates burnout. We believe that the introduction of telemedicine will have an impact on burnout, given the significant change in healthcare provider workflow that it entails.

The need to perform patient care through telemedicine was a significant workflow change for many healthcare providers during the pandemic. Appointment modalities shifted to online formats, both for interactions with patients and communication with other healthcare professionals (14). Appointment modality has extensive effects on provider workflow. Online appointments change the dynamic of communication, the type of care that can be provided, adds an additional layer of logistics, and increase the time providers spend on technology (11). We know that increased use of electronic medical record (EMR) is associated with higher levels of burnout because of increased administrative burdens, decreased patient time, and demands that exacerbate the work-life imbalance (15– 17). Given that telemedicine shares many similarities with EMRs including but not limited to reviewing and documenting patient information, we believe that it might have a similar relationship with burnout rates. Additionally, organizational factors that contribute to burnout including workload expectations, limited opportunities for advancement, diminished social support, and negative leadership behaviors (18), were impacted by the pandemic, and could have been influenced by the introduction of telemedicine into the healthcare workflow. However, recent studies also discuss the importance of workplace flexibility to reduce burnout, which telemedicine may help provide (12,15–17).

While there have been studies on the use and implementation of telemedicine during the pandemic, as well as studies on the prevalence of burnout and stress among healthcare workers during the pandemic, there is very little understanding of the role of telemedicine in contributing to or mediating provider burnout. In summary, it is unclear what effect the introduction and use of telemedicine systems during the pandemic may have had on provider burnout.

In our study, we aim to (1) assess the current state of burnout prevalence among providers who have been using telemedicine to provide care during the pandemic and (2) determine which aspects of telemedicine exacerbate burnout, and which aspects alleviate provider burnout. This study uses the Job Demands-Resources model to categorize the contributing and mediating factors of telemedicine on burnout.

## Background

### Factors Influencing Burnout and Consequences of Burnout

Numerous personal and occupational factors have been found to relate with higher rates of burnout. For example, women in healthcare typically report higher rates of burnout (2,3,12). Specialties that provide frontline care, such as emergency and family medicine, also record higher burnout rates (15,16,18,19). Burnout has a negative effect on patient satisfaction, health outcomes, and access to care as a result of the risk of worker shortages (10,13,15,16,18–20). Burnout has a direct impact on the work life of providers, as employees report decreased job satisfaction and are at a higher risk of depression, substance abuse, and suicide (13,16,19). Unnecessary medical spending on recruitment, training, onboarding, and lost productivity costs have an effect on healthcare cost reduction (13). Burnout has the potential to affect healthcare workers, patients, and organizations, highlighting the urgent need to address burnout and enhance provider well-being. Addressing healthcare burnout is closely aligned with the quadruple aim that many healthcare organizations use to achieve excellence, which provides additional impetus for identifying solutions (21).

### Job Demands-Resources Model

In this study, we classify the telemedicine related factors that contribute to or alleviate burnout through the Job-Demands-Resources (JD-R) Model. The JD-R Model is a framework that classifies job characteristics relating to employee wellbeing either as a job demand, relating to job strain, or resources, relating to motivation (22). Job demands can constitute physical, social, psychological, and organizational components of the job that involve continuous effort; frequently emotional demands, workload, and place of work are considered demands (22). Physical, social, psychological, and organizational components of the job that alleviate demands, assist employees in meeting goals, or provide employees with new opportunities are considered job resources (22).

A key component of this model is the idea that demands and resources each rely on two separate mental processes; demands contribute to the depletion of energy in a worker, while resources contribute to intrinsic and/or extrinsic motivation (22). This model operates under the assumption that job resources can act as a buffer between job strain and employee burnout and examines the direct implications on employee wellbeing (22). This study chose the JD-R model due to its emphasis on employee outcomes and the variety of resources under consideration.

## Methods

### Settings and Participants

This study was conducted in a large, academic medical center in the Midwest. A purposive sample was used for this study. This study recruited from two different worker populations in the hospital who had used telemedicine in the past two years. The first population group was a nurse triage call center that provided care and guidance to patients over telephone calls. The second group consisted of all physicians, nurse practitioners, and physician assistants who had worked in the hospital’s influenza-like illness (ILI) clinics during the previous two years. These populations used telemedicine to provide care to patients with COVID-19 concerns. Students and residents were excluded from this study. Participants were not restricted based on frequency of telemedicine use; the sample included individuals who used telemedicine on a full-time, part-time, and overtime-only basis.

Approval from the Institutional Review Board was granted in August 2022. Following approval, participants were recruited via email invitations sent out from departmental leadership. Two recruitment emails were sent to each group with one week in between. Following the second email, the first author also attended a monthly staff meeting for the ILI clinic providers to provide additional information on the study and invite staff to participate. No similar departmental meeting was available to recruit from the triage call center. Participant documentation of consent was waived to ensure confidentiality of data.

### Experimental Procedure

A Qualtrics survey was emailed to the participant population through department leadership. Seven questions captured respondent characteristics such as gender, ethnicity, age, and years in their field. The remainder of the survey used the validated Maslach Burnout Inventory assessment tool (12). The MBI is considered a ‘gold standard’ assessment tool because it can discern the general experience of exhaustion from burnout by incorporating an occupational context and assessing different dimensions that can influence one’s mental state (23). The three dimensions of burnout under the MBI are: emotional exhaustion (EE), depersonalization (DP), and personal accomplishment (PA) (12). Numerous studies have demonstrated its reliability and validity (12,24).

The MBI consists of 22 statements and a Likert scale on the frequency of agreement. For example, a ‘0’ indicates that the respondent has never felt what the statement suggests, while a ‘6’ indicates that the respondent agrees with that statement every day. Participants were initially informed that the study’s purpose was to assess job attitudes. Per recommendations of the MBI manual, the true intent of the study, the evaluation of burnout, was not initially disclosed to the participants, to prevent reactive emotions from influencing their responses (12). The survey link was live for one month and participants could complete the survey in multiple sittings if they preferred. Once the survey was completed, participants were debriefed on the true nature of the study, provided with information on techniques to mediate burnout, and offered the opportunity to participate in follow-up interviews.

The first author scheduled a Zoom follow-up interview with survey participants who were willing to participate in interviews. The interview consisted of nine questions and lasted between 15-30 minutes. Follow-up interviews explored participants’ positive and negative experiences with telemedicine. Participants described how they used telemedicine to provide patient care. Interviews also gathered participants’ ideas on how to better support this technology in patient care. The audio was recorded using Audacity (25) software and then manually transcribed by the first author.

### Analysis Procedure

Respondent demographics were analyzed using descriptive statistics. Burnout was classified by determining a prevalence rate and generating burnout profiles of the respondents. Burnout prevalence was determined using the cut-off values of >= 27 for EE total score or >= 10 for DP total score (13). This method can help differentiate between burnt-out and non-burnt-out individuals (24). Burnout profiles were generated by assessing low, moderate, and high designations in each burnout dimension. These classifications were determined by comparing responses to normative data from the MBI 4^th^ Edition Manual (12). Correlation coefficients were calculated to understand the relationships between burnout dimension scores and respondent characteristics, such as age and reported months of telemedicine use.

Interview data was coded by the first author using MAXQDA (26). The codebook was developed based on the JD-R model components and current evidence from literature, but was modified as additional demands and resources were discussed by interview participants. Participant responses were coded as either a specific demand or resource. For each coded segment, it was further classified as either: + improving the demand or resource, -worsening the demand or resource. Demands are aspects of the work that require continuous effort, and resources are aspects of the job that alleviate a demand or fuels worker motivation (22). An increase in a demand or a reduction in a resource is considered a contributing factor to burnout. A decrease in a demand or an increase in a resource is considered a mediating factor of burnout. The resulting codes and coded segments were then analyzed for emerging themes.

## Results

### Survey Findings

#### Results and Participants

The overall survey response rate was 34% (32/94), and nine of the 32 respondents (28%) completed interviews. All respondents were from the urgent care ILI clinic population. Respondent characteristics and findings are displayed in tables 1, 2, and 3. Most (91%, n = 29) of the respondents were female. More than half (59%, n = 19) of the respondents were nurse practitioners. All respondents were white.

**Table 1:**
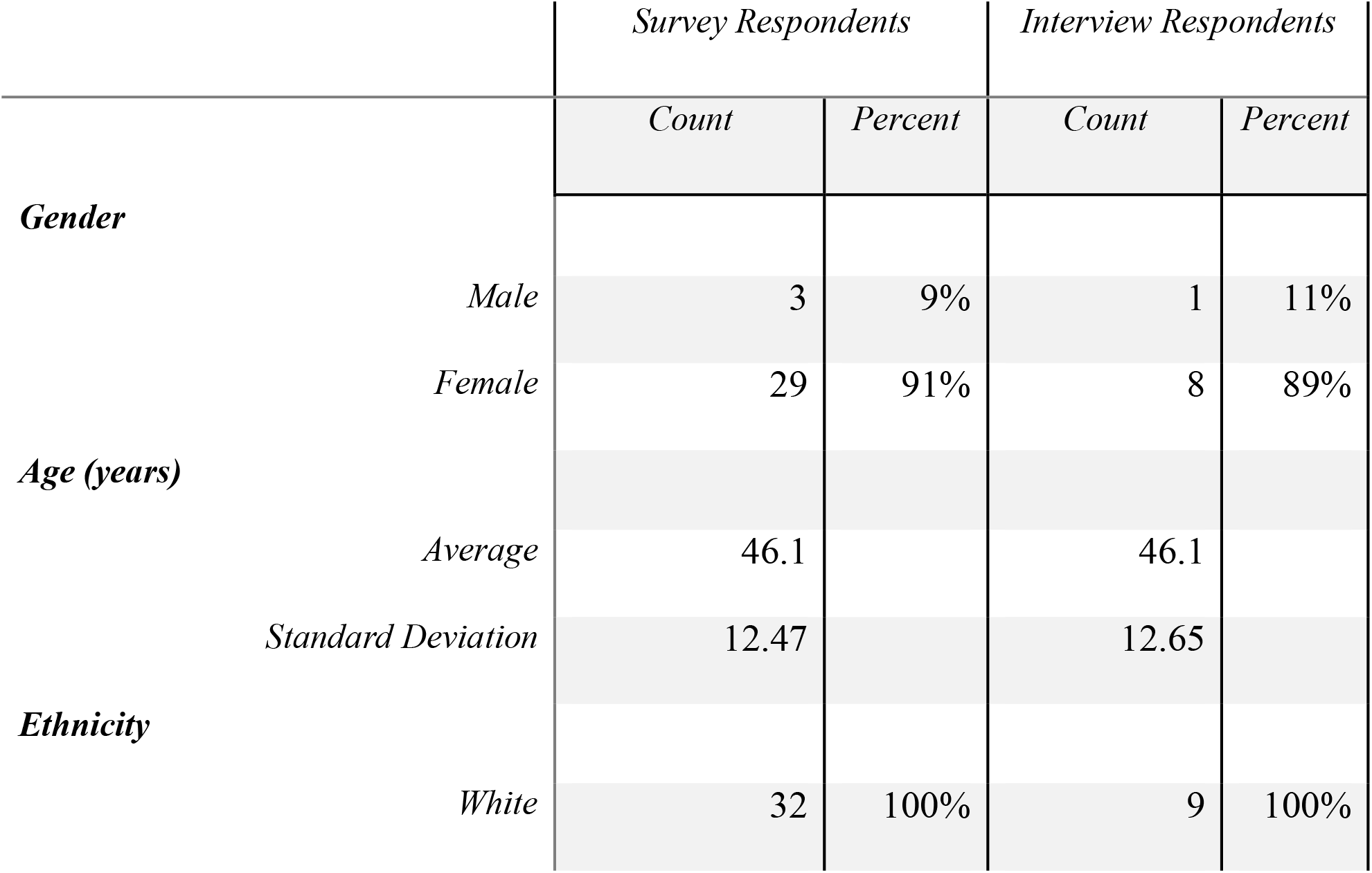
Survey and Interview Respondent Demographics

|  | *Survey Respondents* |  | *Interview Respondents* |  |
| --- | --- | --- | --- | --- |
|  | *Count* | *Percent* | *Count* | *Percent* |
| ***Gender*** |  |  |  |  |
| *Male* | 3 | 9% | 1 | 11% |
| *Female* | 29 | 91% | 8 | 89% |
| ***Age (years)*** |  |  |  |  |
| *Average* | 46.1 |  | 46.1 |  |
| *Standard Deviation* | 12.47 |  | 12.65 |  |
| ***Ethnicity*** |  |  |  |  |
| *White* | 32 | 100% | 9 | 100% |

**Table 2:** Participant Professional Characteristics

|  | *Survey Respondents* |  | *Interview Respondents* |  |
| --- | --- | --- | --- | --- |
|  | *Count* | *Percent* | *Count* | *Percent* |
| ***Professional Role*** |  |  |  |  |
| *Nurse Practitioner* | 19 | 59% | 6 | 67% |
| *Physician* | 8 | 25% | 0 | 0% |
| *Physician Assistant* | 5 | 16% | 3 | 33% |
| ***Years in Field*** |  |  |  |  |
| *Average* | 10.73 |  | 5.56 |  |
| *Standard Deviation* | 9.03 |  | 4.07 |  |
| ***Months of Telemedicine Use*** |  |  |  |  |
| *Average* | 19.10 |  | 21.88 |  |
| *Standard Deviation* | 9.31 |  | 9.06 |  |
| ***Work Assignment to Telemedicine*** |  |  |  |  |
| *Originally Worked* | 16 | 50% | 5 | 56% |
| *Reassigned* | 10 | 31% | 1 | 11% |
| *Volunteered Overtime* | 6 | 19% | 3 | 33% |

**Table 3:**
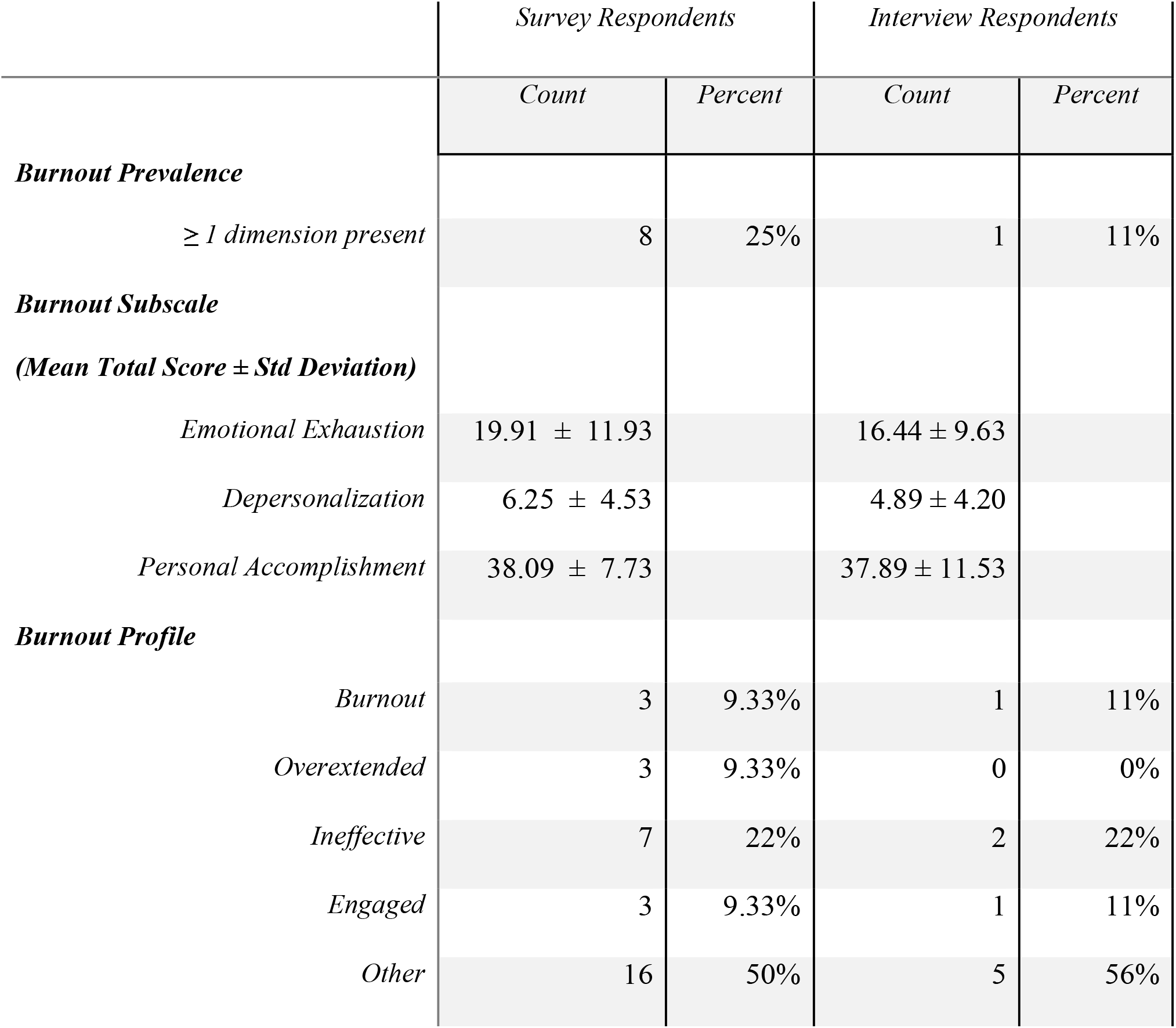
Respondent Burnout Scores

#### Burnout Prevalence

Within this sample, 25% (n = 8) of the respondents showed at least one dimension. There was no statistically significant difference of prevalence across gender, professional role, or work assignment to telemedicine. Burnout profiles (12) indicated the following: Seven individuals (22%) were found to be ‘Ineffective’; this grouping is defined by low PA scores with low or moderate EE and DP scores. Three respondents (9.3%) were classified as ‘Burnout’; this means that respondents had high scores of EE and DP and low PA. Three respondents (9.3%) were labeled ‘Overextended’, meaning that respondents had high EE and moderate scores of DP and PA. Three respondents (9.3%) were classified as ‘Engaged’; this means that these respondents had high PA, low EE, and low DP scores. All other respondents (50%) did not fit a pre-defined burnout profile defined by the Maslach Burnout Inventory (12).

### Burnout Dimension Correlations

Correlations were calculated for each of the three burnout dimensions with respondent age, years in the field, and reported months of telemedicine use. Note that months of telemedicine use does not differentiate employees who used telemedicine full time, part time, or only when volunteering overtime; only the start of telemedicine use was referenced in the question. There was a negative correlation of R = -0.3769 (*p = 0*.*0439*) between total personal accomplishment scores (assessed by the MBI) and reported months of telemedicine use. This is shown in Figure 1.

**Figure 1:**
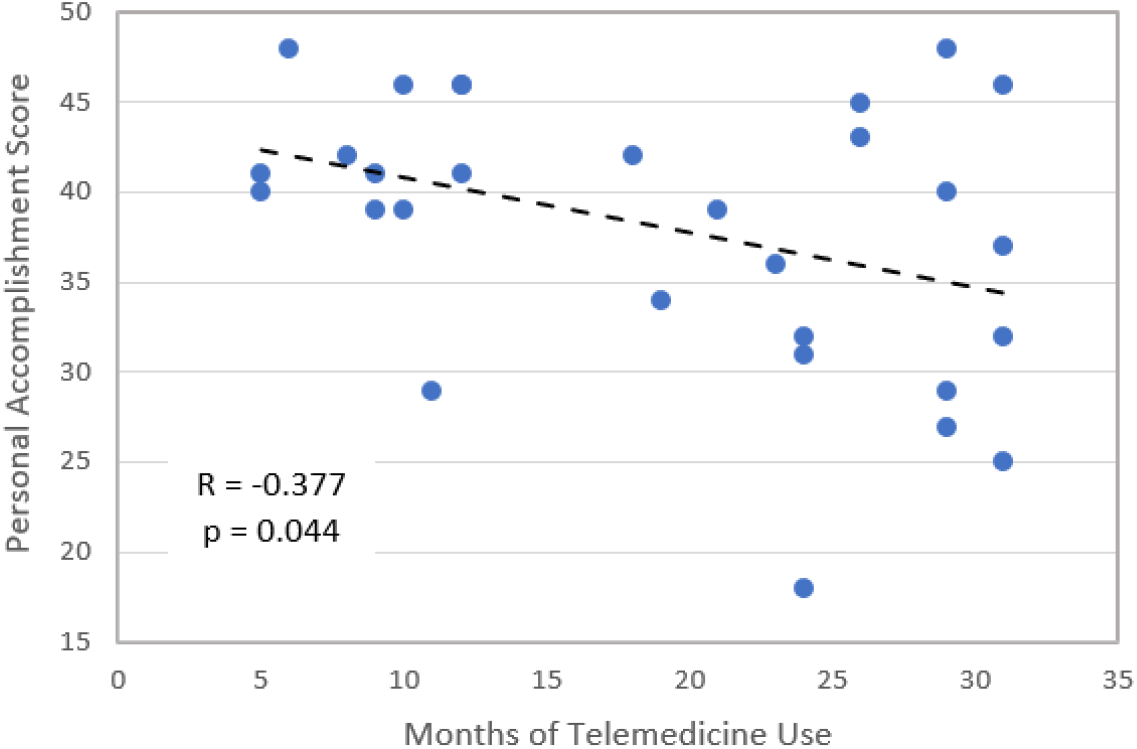
Scatterplot of Personal Accomplishment Scores and Months of Telemedicine Use

### Interview Findings

#### Frequently Mentioned Contributing and Mediating Factors

The top three mentioned contributing and mediating factors are listed in tables 4 and 5. These tables also include quotes from the interviews to demonstrate how the demands and resources apply in context. The top contributing factors indicated demands that were worsened for providers due to telemedicine; the top mediating factors highlighted resources that were improved for providers.

**Table 4:** Most Mentioned Contributing Factors

| Contributing Factor | Underlying Mechanism | Number of Mentions | Quote |
| --- | --- | --- | --- |
| - Difficult Patient Cases | Added demand | 12 | “There are limitations of being able to really help people when it seems like they are really lost in the healthcare system overall. Like people who never establish care and are wanting things that a primary care provider should be doing and following up with. And that can be frustrating when you can tell that these people constantly come to these telemed or urgent care centers and it’s like ‘You’ve gotta get somebody because we can’t be doing what we are doing.’ This isn’t the right setting.” |
| - Technology Stress | Added demand | 9 | “The frustration, sometimes technology is just technology. Where you’ll have bad Wi-Fi connection or you’re trying to reach out to them because they can’t figure out their mic, so you have to physically call them and then they have your phone number.” |
| - Time Pressure | Added demand | 8 | “... The facility pushes for short appointment times, and if you are unable to complete the appointment in the amount of time, you can get behind for the rest of the day. If this happens with 2 or 3 patients in a row, it can set you behind a whole hour. I know [here] we have been behind 2 or 3 hours before because of this.” |

**Table 5:** Most Mentioned Mediating Factors

| Mediating Factor | Underlying Mechanism | Number of Mentions | Quotes |
| --- | --- | --- | --- |
| + Patient Experience | Added resource | 11 | “I feel like patients were a little happier with the telehealth as well. So, when you start with somebody that is a little happier in their visit, yaknow, they didn’t have to worry about parking and they didn’t have to worry about the drive in, they aren’t worried about the weather, that’s a huge stress relief for them. So that makes their baseline happiness a little better, which makes everything go that much better.” |
| + Remote Work | Added Resource | 7 | “I was burning out and Covid exacerbated all the things that were contributing to burnout for me. And the telemedicine change was a nice change. It was a switch getting to work from home because I have never, obviously in healthcare you can’t really work<br>*from home in certain roles. But it was really nice to be able to work from home.”* |
| +<br>Technology<br>Stress | Improved<br>Demand | 5 | • *“The organization supports it well, they provide us with the equipment that we need, they give us training, and there are always people and resources there to help support us using telemedicine.”* |

## Discussion

One of the most notable findings from this study is the low prevalence of burnout in the sample of providers who use telemedicine to provide care to patients. Only 25% of the respondents (n=8) in this study reported at least one dimension of burnout. Before the pandemic, reported levels of burnout in the healthcare industry were around 40-54% (13); post-pandemic studies indicate that the prevalence has risen to as high as 62% (2). However, the burnout rates in this study do not reflect this trend. First, both the pre-pandemic and post-pandemic studies on clinician burnout do not differentiate burnout rates of telemedicine providers from burnout rates of other clinicians, so at present we do not know whether and to what extent telemedicine contributes to the high rates of burnout reported in the literature. Second, our findings also suggest that telemedicine has the potential to mediate clinician burnout. This mediation of burnout could be possible because the benefits of telemedicine have a greater impact on clinician wellbeing than the drawbacks.

Moreover, by juxtaposing correlation findings with trends in burnout profiles, we identified a possible explanation for burnout among telemedicine providers. The “telemedicine use in months” and scores on “personal accomplishment” had a statistically significant negative correlation of R = -0.3769 (*p = 0*.*0439*) indicating that clinicians who mention using telemedicine for longer periods of time typically report feeling less accomplished in their professional work. Additionally, the most reported burnout profile, besides ‘Other’, was ‘Ineffective.’ This profile group is distinguished by its low personal accomplishment scores. These complementary findings suggest that burnout manifests in this setting by diminishing feelings of personal accomplishment. This relationship is expected, based on studies that demonstrate increased feelings of underachievement when participating in digital meetings compared to in-person visits (11). Digital patient appointments may make providers feel less satisfied or inadequate at work, which could amplify burnout manifestations. Other studies have also shown that the more frequent virtual meetings are, the more fatigue that can arise from using these technologies (27), which would help explain how the total amount of time a provider uses telemedicine impacts their feelings of accomplishment.

Lastly, half of the participants did not fit into an MBI Burnout Profile when classifying respondents into ‘Low,’ ‘Moderate,’ and ‘High’ categories in each burnout dimension. These profiles are a newer method for understanding burnout. The majority of burnout profiles require a ‘High’ or a ‘Low’ score in burnout dimensions to fit into a category (12). In this study, a large proportion of respondents reported moderate attitudes in various dimensions. There is a need to better understand these profiles and differentiate between moderate experiences across multiple dimensions in order to capture nuanced workplace experiences, particularly for workers who may not yet be at an extreme in a certain burnout dimension. Identifying and understanding the moderate burnout profiles will also allow us to intervene early before moderate burnout becomes severe.

### Contributing Factors of Telemedicine on Provider Burnout

The factors that contribute to provider burnout (Figure 2) seem to stem from constraints inherent to telemedicine technology (i.e., inappropriate use of telemedicine, administrative burden, technology issues, isolation with remote tools, etc.), as well as elements that are influenced by how an organization implements telemedicine (i.e., time pressure, appointment scheduling options, workflow issues, training for providers and patients on the technology).

**Figure 2:**
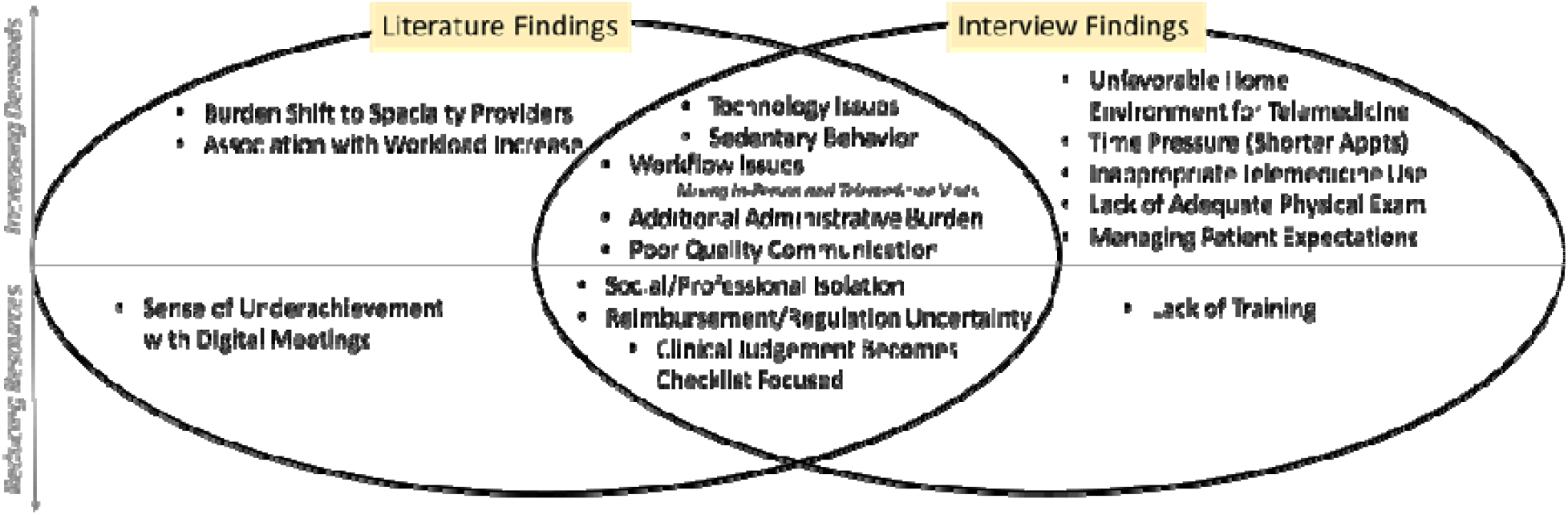
Contributing Factors of Telemedicine on Provider Burnout

Consistent with the literature, our findings indicate that characteristics inherent to telemedicine technology may contribute to the burnout of healthcare workers. One of the most reported contributing factors, mentioned in six of the nine interviews in our study, was the added demand from inappropriate use of telemedicine; that is, appointments that are scheduled as virtual visits when they should have been scheduled as in-person visits. This demand is inherent to the technology, as not all patient care can be performed virtually, and not all automated scheduling screenings can detect inappropriate visits. Literature, however, spoke little of this as an added demand on providers, and only discussed inappropriate telemedicine use in reference to which departments would least benefit from the use of telemedicine (17).

One demand that is exacerbated by the use of telemedicine is the increased administrative burden the user faces (4,15,17,28,29). This can include tasks from in-person appointments that are reassigned to the provider during telemedicine visits, as well as additional charting and documentation tasks required for telemedicine appointments. Another added demand is technology shortcomings (13,15,30); this includes hardware and software issues experienced by providers and/or patients. Our study findings also revealed the provider’s perception of inadequacy to provide troubleshooting support to the many technology issues experienced by the patient. The patient’s struggle with telemedicine technology was a factor that was not previously reported in the literature. This suggests that user experience efforts should also consider the patient perspective, and not just that of the provider. In addition, our findings indicate that providers face social and professional isolation when providing care via telemedicine, confirming previous studies that highlight the inadequacy of virtual visits in fostering the same face-to-face connection and personal relationships that in-person visits facilitate (31,32).

One of the most surprising findings was that multiple articles speculated that telemedicine would increase rates of burnout by contributing to a work-life imbalance (3,32,33), but none of the interviewees in our study mentioned this. In fact, multiple interviewees cited an improvement in work-life balance. We know from other studies that negative work-life balance is associated with higher rates of burnout and positive work-life balance is correlated with well-being (34,35). The interview responses in this study suggest that telemedicine likely promotes a good work-life balance, despite literature suggesting that it may worsen this balance.

### Mediating Factors of Telemedicine on Provider Burnout

Literature findings are generally consistent with frontline reports (Figure 3) on the benefits of telemedicine, which we think has the potential to alleviate burnout. Our findings concur with previous research showing telemedicine’s potential to reduce the hazard of exposure to illnesses (3,28,33,36), one of the foremost reasons to implement telemedicine during the pandemic.

**Figure 3:**
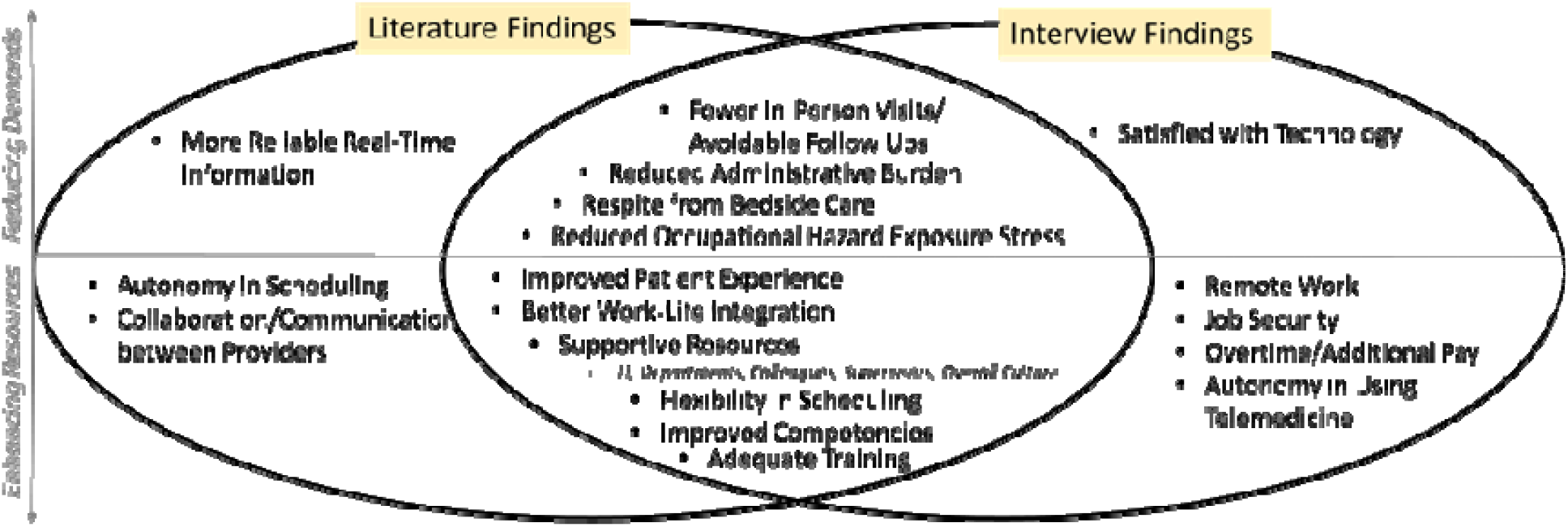
Mediating Factors of Telemedicine on Provider Burnout

Our findings also confirm that the reward of improved patient experience, better work-life integration, and improved competencies are key benefits of telemedicine (3,13,17,28,31,33,36,37). Because patients were satisfied with the telemedicine option, providers could feel rewarded. Providers were also proud of the far-reaching impact they could have by offering remote patient care services. The remote aspect of telemedicine allowed for better work-life integration by reducing commute times, freeing up more time for providers’ families and hobbies, and giving providers with children greater flexibility. Telemedicine also provided additional competency opportunities for providers to learn new methods of providing care, familiarize themselves with new technology, and practice in settings outside of their traditional scope. All these resources motivated the providers and/or enabled them to continue to focus on providing care.

Studies report that one of the key benefits of telemedicine is its ability to improve autonomy by giving providers more control over their schedule (3,13,31,33,36–38). In our interviews, autonomy was not discussed in relation to scheduling, but in reference to appointment modality. Eight of the nine interviewees did not mention autonomy at all; but one interviewee stated that providers should be able to choose between telemedicine or in-person appointments, pointing out the need for autonomy in appointment modality. The interviewee expressed, “I knew going in [to this position] that I was going to be doing telemedicine, but I think several [other staff members] had it added to their job description and weren’t happy about it… If providers want to do it, that’s great. But forcing providers who don’t want to do it, that’s not good.” This finding is interesting because the employing organization has the potential to influence how telemedicine can benefit employee’s perceived autonomy. When an organization implements a telemedicine program an organization has the choice to determine how providers set their schedules, where they can work from and how they choose to practice. The degree of control that an organization grants its providers over how they structure their work has a direct impact on perceived autonomy and may influence providers’ motivation to perform the work.

In summary, the pandemic has only exacerbated the growing concerns regarding healthcare worker burnout. But telemedicine technology holds promise if, prior to implementation efforts, we systematically understand the impact of technology on healthcare worker well-being. Understanding the contributing and mediating factors of telemedicine to burnout can help identify interventions that work at the technology level, which can be initiated by developers, and interventions at the work organization level, which can be initiated by organizational leaders.

## Limitations

This study only examines the telemedicine experience of a single institution and a single platform, which limits the technology-specific factors and recommendations that could be derived from a study of multiple platforms. Nonetheless, using the JD-R lens in conjunction with the burnout inventory is a crucial first step in identifying contributors and mediators of burnout from the perspective of occupational well-being.

This study’s survey sample size was small, primarily due to pandemic-related workload constraints. When analyzing burnout, the amount of telemedicine use was not quantified as an exposure. Therefore, this study cannot infer a causal relationship and does not provide a dose-response analysis of telemedicine use and burnout. However, our study addressed the need for a deeper understanding of burnout. In addition, “months of telemedicine use” does not capture frequency of use, as our objective was to identify how long an individual has been using the system, not how frequently. We acknowledge that, in addition to duration, frequency may also have an impact on burnout, and this should be considered in future research. We recognize the possibility of survival bias, as burnt-out employees may have already left the profession.

## Recommendations

Based on contributing and mediating factors identified in this study, we suggest possible opportunities for improvement and intervention that can alleviate burnout by reducing demands or enhancing resources (Table 6). To combat low personal accomplishment in online work settings and its resulting impact on burnout, one could redesign technology or provide organizational support to reward and stimulate achievement.

**Table 6:** Design Implications for Telemedicine

|  | Alleviates Demand | Enhances Resource |
| --- | --- | --- |
| Technology<br>Level | • Improve automated screening for appropriate visits ahead of time<br>• Automated communication with patients upon scheduling regarding the functional limitations of the technology in order to manage expectations | • Staff interface modifications and training<br>○ User interface design to simulation achievement/accomplishment<br>○ Training on hardware, software, patient troubleshooting, and skills for conducting a visit<br>• Patient interface modifications and training |
| Work Organization<br>Level | • Additional staff assigned to connect with patient beforehand and check their technology, connection, review medications/allergies/etc.<br>• Block for longer visits, especially when staff are first starting with telemedicine<br>• Finding ways to integrate telemedicine to address more patient conditions<br>○ Alleviate demands on providers in areas where telemedicine is less feasible | • Improve organizational resources for technology and office supplies to work from home<br>• Organizations allow the provider autonomy in offering telemedicine services<br>○ Potential financial incentives to offer services<br>• Organizational ways to acknowledge and recognize telemedicine providers to intervene on low personal accomplishment sentiments |

### Technology Level Recommendations

Design changes to the technology will benefit the end user by alleviating stressors or improving motivation. For instance, using telemedicine for visits that are more appropriate for an in-person visit was one of the most difficult challenges faced by clinicians. To reduce this demand, automated screening for appropriate visits could be improved to more accurately capture and integrate the reason for the visit when scheduling the appointment.

Participants noted that automating the communication about technology limitations to patients will also help set patients’ expectations and better prepare them for telemedicine visits. This may reduce the troubleshooting demands for clinicians during telemedicine sessions. Additionally, training resources for both staff and patients can be improved. Staff training should address hardware set-up, software training, and patient troubleshooting. Readily available patient training would provide both patients and providers with resources for solving common technology issues.

Literature and study findings both highlight the sense of under accomplishment with virtual tasks, which may be addressed with user interface design modifications to simulate achievement (11) and potentially increase motivation.

### Work Organization-Level Recommendations

Organizations themselves can have a positive or negative impact on the demands placed on employees. One demand that organizations can control is support staff to help start appointments by ‘rooming the patient’, checking connectivity, and reviewing charts. These support staff are available for in-person visits, but many interviewees noted that they found themselves taking on these tasks in telemedicine settings. The demand of time pressure can be alleviated by increasing appointment duration, particularly for providers who are new to telemedicine. Additionally, expanding telemedicine to other areas can ease overall healthcare system demand. Interviewees mentioned that integrating telemedicine into more specialties can improve management of chronic diseases, thereby preventing and reducing emergency visits and more appointments downstream. Making appointments more accessible can help with patient compliance and allow busier departments to offset less urgent cases.

Organizations can also increase resources for providers. One example would be ensuring providers have the necessary technology and office equipment to work from home. Also, organizations can prevent employee burnout by allowing providers to determine where, when, and how providers set their schedules. Furthermore, the interviewees noted that improved financial incentives for providers who are willing to use telemedicine can help motivate providers to consider integrating these technologies into their practice. Lastly, to proactively combat feelings of low personal accomplishment, organizations can find ways to recognize and acknowledge employees in telemedicine and remote work settings.

### Future Research Recommendations

Future research can quantify telemedicine use and burnout manifestations to establish causality and examine the effectiveness of telemedicine as an intervention method for burnt out individuals. More research is also needed to help differentiate between moderate scores in burnout profiles and to implement preventive measures in place for employees who have not yet reached extreme levels of burnout.

### Conclusion

The burnout crisis has been a prevalent and growing issue in the healthcare industry, with the COVID-19 pandemic impacting various aspects of provider work. This study investigated the role of telemedicine in clinician burnout and found a lower than expected prevalence of burnout (25%) among providers that have used telemedicine. Nevertheless, this study found that telemedicine providers can experience burnout if their sense of personal accomplishment is undermined. Contributing and mediating factors were identified and categorized to determine interventions and improvements that technology designers, hospital administrators, and researchers can have an impact on.

## Data Availability

De-identified data produced in the present study are available upon reasonable request to the authors.

## Acknowledgements

I would like to thank Laura Cullen, Associate Research Scientist, Dr. Katie Imborek, MD, and Robert Linnell, PA-C, for serving as points of contact and assisting with recruiting participants. Laura also served on my thesis committee. I would also like to thank my advisor, Dr. Priyadarshini Pennathur, for her continued support and guidance throughout my time at the University of Iowa. I would also like to thank Dr. Nathan Fethke and Dr. Arunkumar Pennathur for their feedback and support as members of my thesis committee.

Thanks to Laura Cullen, Dr. Katie Imborek, and Robert Linnell for assisting with the study. Thanks, also, to my thesis committee for their support and guidance: Dr. Priyadarshini Pennathur, Dr. Nathan Fethke, Dr. Arunkumar Pennathur, and Laura Cullen.

Valerie Boksa was supported by grant #2128495 from the National Science Foundation (NSF) during the conduct of this work. Valerie was also supported by the National Institute for Occupational Safety and Health (NIOSH) training grant #T42OH008491 through the Heartland Center for Occupational Safety and Health at the University of Iowa. The findings of this study are those of the authors and do not represent the views of NSF and NIOSH.

